# Glucagon-like peptide-1 receptor agonists modestly reduced blood pressure among patients with and without diabetes mellitus: A meta-analysis and meta-regression

**DOI:** 10.1101/2024.01.29.24301971

**Authors:** Frederick Berro Rivera, Grace Nooriza O. Lumbang, Danielle Rose Magno Gaid, Linnaeus Louisse A. Cruz, John Vincent Magalong, Nathan Ross B. Bantayan, Kyla M. Lara-Breitinger, Martha Gulati, George Bakris

## Abstract

**Background:** The cardiovascular benefits provided by glucagon-like peptide-1 receptor agonists (GLP-1RAs) extend beyond weight reduction and glycemic control. One possible mechanism may relate to blood pressure (BP) reduction. We aim to quantify the BP lowering effect by GLP1-RAs.

**Methods:** A comprehensive database search for placebo-controlled randomized controlled trials (RCTs) on GLP-1RA treatment was conducted until December 2023. Data extraction and quality assessment were carried out, employing a robust statistical analysis using a random effects model to determine outcomes with mean difference (MD) in millimeters mercury (mmHg) and 95% confidence intervals (CIs). The primary endpoint was the mean difference in systolic and diastolic BP. Subgroup analyses and meta-regression were done to account for covariates.

**Results:** Compared to placebo, GLP-1RAs modestly reduced SBP (semaglutide: MD −3.40, [95% CI −4.22 to −2.59, p<0.001], liraglutide: MD −2.61, [95% CI −3.48 to −1.74, p<0.001], dulaglutide: MD −1.46, [95% CI −2.20 to −0.72, p<0.001] and exenatide: MD −3.36, [95% CI - 3.63 to −3.10, p<0.001]). This benefit consistently increased with longer treatment duration. Established people with type 2 diabetes experienced less SBP lowering with semaglutide. DBP reduction was only significant in the exenatide group (MD −0.94, [95% CI −1.78 to −0.1], p=0.03). Among semaglutide cohorts, mean change in hemoglobin A1c and mean change in body mass index were directly associated with SBP reduction.

**Conclusion:** Patients on GLP-1RA experienced modest SBP lowering compared to placebo. Only exenatide reduced DBP. Further studies are needed to clarify the mechanisms and the clinical benefit of GLP-1RA effects in BP reduction.

## INTRODUCTION

In recent years, glucagon-like peptide-1 receptor agonists (GLP-1RAs) have emerged as a first-line drug in the treatment of type 2 diabetes mellitus (T2DM).^1^ GLP-1RAs have also been found to significantly reduce the risk of major adverse cardiovascular events (MACE) in patients with T2DM, as demonstrated by multiple large cardiovascular outcome trials (CVOTs).^2^ Recently, the STEP-HFpEF (Semaglutide in Patients with Heart Failure with Preserved Ejection Fraction and Obesity) and SELECT (Semaglutide Effects on Cardiovascular Outcomes in People With Overweight or Obesity) trials, found that semaglutide led to improvements in symptoms, physical limitations, and exercise function in patients with heart failure, as well as decreased the risk of MACE in overweight and obese patients.^3, 4^ The available evidence demonstrates the significance of GLP-1 receptor agonists in the treatment of obesity and the reduction of cardiovascular risk in patients with type 2 diabetes mellitus.^5, 6^ The cardiovascular benefits provided by GLP-1RAs are found to be significant, independent of its effects on glycemic control.^7^ However, the specific mechanisms underlying the cardiovascular protection provided by GLP-1RAs have not yet been fully elucidated. One such pathway may be the reduction of blood pressure, a well-known CV risk factor.^8, 9^ GLP-1 receptors are found in the hypothalamus and brainstem, the heart and vasculature, and the kidneys, suggesting multiple potential mechanisms that underlie the blood pressure-lowering effects of GLP1-RAs, independent of their effect on glycemic control.^10, 11^ Among these include, but are not limited to, inhibition of sympathetic nerve excitation, reduction of vascular inflammation and arterial stiffness, and promotion of natriuresis and diuresis.^11^ Previous studies have shown that GLP-1RAs reduce blood pressure by as much as 2-3 mmHg, with systolic BP being lowered more than diastolic BP.^12, 13^ In subsequent meta-analyses, liraglutide and semaglutide have been demonstrated to provide a significant reduction in systolic blood pressure.^14, 15^ Nevertheless, the significance of the effect on BP lowering by GLP1-RAs overall and across different subgroups remains inconclusive, and thus there is a need to bridge existing knowledge gaps with the latest evidence from recent trials in GLP-1RA use.

In this study, we conducted a thorough systematic review and meta-analysis of RCTs on the four widely used GLP-1RAs (Semaglutide, Liraglutide, Exenatide and Dulaglutide). We aim to determine the efficacy of GLP-1RA in reducing SBP and DBP across different GLP-1RAs. Subgroups based on dose, treatment duration, baseline mean SBP and DM status, as well as regression analyses were done to account for covariates.

## METHODS

This study was reported under the Preferred Reporting Items for a Review and Meta-Analysis (PRISMA)^16^, and the checklist^17^ was followed **(Figure S1).** Certainty of evidence was rated using the Grades of Recommendation, Assessment, Development, and Evaluation (GRADE) framework.^18^ This study was registered in the International Prospective Register of Systematic Reviews (PROSPERO)^19^, with the identification number CRD42024497765.

### Data Sources and Searches

The literature search was performed using PubMed/MEDLINE, Ovid/Embase, and clinicaltrial.gov databases from database inception until December 2023. Search terms included “glucagon-like peptide-1 receptor agonists”, “GLP-1 agonist”, “GLP-1RA”, “semaglutide”, “dulaglutide”, “exenatide”, “liraglutide”, “placebo”, “blood pressure”, “systolic blood pressure” “diastolic blood pressure”, “randomization”, “clinical trials”, “intervention studies” and synonyms. Citations of selected articles and any relevant studies that evaluated GLP-1RA and blood pressure reduction were reviewed. After removing duplicates, records were reviewed at the title and abstract level, followed by the screening of full text based on our study criteria.

### Study Selection

Eligible phase II or phase III, double-blind randomized controlled trials (RCTs) comparing treatment with GLP-1RA with placebo in adult patients aged 18 years and above were included. Moreover, the studies must have reported baseline SBP and DBP and blood pressure reduction in weighted mean difference. Studies must have reported the mean change in weight, mean change in hemoglobin A1c, and mean change in body mass index. Furthermore, studies must have data on the baseline mean SBP and DBP and must mention the diabetes status of the cohort. Studies were excluded if (1) they did not report a control arm, (2) there was an active comparator, and (3) mean change was not reported. Other agents such as albiglutide, lixisenatide, efpeglenatide, were excluded because cardiovascular outcomes are not well established with these agents. We also excluded RCTs with participants younger than 18, and those reporting interim or post hoc analysis. Cross-over trials were also excluded due to the nature of the outcomes considered. Review articles, case reports, letters to the editor, commentaries, proceedings, laboratory studies, and other non-relevant studies were excluded as well.

### Data Extraction

Key participant and intervention characteristics and reported data on efficacy outcomes were extracted independently by two investigators (GOL and DMG) using standard data extraction templates. Any disagreements were resolved by discussion or, if required, by a third author (FBR). Data on the following variables were extracted: first author’s name, year of publication, journal, study phase, interventional and control treatments, randomization method, analysis tool, number of randomized patients, and demographic and clinical data (e.g., age, sex). In case of uncertainties regarding the study data, we contacted the authors of the specific study for additional information. Quality assessment was performed independently by two review authors using the Revised Cochrane risk-of-bias tool for randomized trials.

### Outcome Measures

The primary endpoint of this meta-analysis and meta-regression was the weighted mean difference (MD) in SBP and DBP. Additionally, subgroup analyses were performed for applicable studies on the (1) duration of treatment in weeks, (2) dose, (3) mode of administration, (4) mean baseline SBP, and (5) DM status of the cohorts.

### Bias Assessment

All included studies reported a central randomization process, and outcomes were objectively determined. The included studies reported all primary and secondary outcomes as pre-specified in their protocols, so the risk of bias for selective reporting was judged as low. Three authors (JM, DMG, and GOL) independently assessed the risk of bias based on the Cochrane Risk of Bias Tool **(Figures S2)** for studies that fulfilled the inclusion criteria. Disagreements between the reviewers were resolved by consensus.

### Statistical Analysis

We pooled all estimates using a random effects model based on the restricted maximum likelihood (REML) model. Effect sizes were expressed using mean differences with 95% confidence intervals (CIs). Funnel plot and Egger test were used for estimation of publication bias. For all outcomes, the significance level was set at a p-value of <0.05 or 95% CI not including 1. Both Cochran’s Q and Higgins and Thompson’s I2 statistics were generated to describe the heterogeneities among the studies. We calculated the *I*^2^ statistics (0–100%) to explain the between-study heterogeneity, with *I*^2^≤25% suggesting acceptable homogeneity, 25% < *I*^2^≤75% suggesting moderate heterogeneity, and *I*^2^>75% suggesting high heterogeneity. Forest plots were used to plot the effect size, either for each study or overall. Publication bias was evaluated by graphical inspection of funnel plot; estimation of publication bias was quantified by means of Egger linear regression test. Meta-regression analysis was performed, reporting results as *P* value. Stata version 18 (*StataCorp, College Station, TX*) was used to conduct the included studies’ meta-analyses and meta-regression.

## RESULTS

A literature search through December 2023, yielded 4,178 potentially relevant references on GLP-1RA. (**Figure S1**). Of these, 1,156 duplicates were removed. A total of 1,221 studies with unrelated interventions, outcomes, populations, non-original data (e.g., meta-analysis or review), descriptive or observational study design, and study protocols were excluded. 635 articles were removed for not meeting the eligibility criteria. The remaining 66 related studies were retrieved as full-text publications for detailed evaluation. Finally, 63 studies were included in the final meta-analysis. 53,072 participants were included, 32, 113 (60.51%) of which are females. The rest of study characteristics are shown in **Table 1**. Majority of the RCTs were multinational and sponsored by drug companies.

**Table 1:** Characteristics of included studies.

| Author (year) /<br>Name of Trial | Phase | Population | No. of<br>patients | Age, years<br>(mean) | Female, n<br>(%) | Drug (dose and<br>frequency) | Comparator | Duration of<br>treatment (weeks) | Primary endpoint |
| --- | --- | --- | --- | --- | --- | --- | --- | --- | --- |
| **Jorsal et al.**<br>**(2016) / LIVE**<br>**Trial<sup>20</sup>** | Phase 2 | Patients with or without<br>diabetes who had CHF, an<br>LVEF ≤45% and NYHA<br>functional class I–III | 241 | 65 | 26 (10.8) | Liraglutide (1.8 mg<br>OD) | Placebo | 24 weeks | Change in LVEF |
| **Loganathan et**<br>**al. (2022) /**<br>**ELAID Trial<sup>21</sup>** | Phase 4 | Patients with T2DM and<br>without concurrent<br>antiplatelet therapy | 16 | 60 | 6 (37.5) | Liraglutide (1.8 mg<br>OD) | Placebo | 26 weeks | Change in maximum slope<br>of platelet aggregation |
| **Kadowaki et al**<br>**(2022) / STEP 6**<br>**Trial<sup>51</sup>** | Phase 3 | Adults with at least one self-reported unsuccessful<br>dietary attempt to lose body<br>weight | 401 | 51 | 148<br>(36.9) | Semaglutide (2.4 mg<br>and 1.7 mg QW) | Placebo | 68 weeks | Change in BW and<br>proportion of subjects<br>achieving at least 5%<br>reduction of baseline BW |
| **Davies et al.**<br>**(2021) / STEP 2**<br>**Trial<sup>52</sup>** | Phase 3 | Adults with T2DM and with<br>at least one unsuccessful<br>dietary effort to lose weight | 807 | 55 | 413<br>(51.2) | Semaglutide (2.4 mg<br>QW) | Placebo | 68 weeks | Percentage change in BW<br>from baseline and loss of at<br>least 5% of baseline BW |
| **Gadde et al.**<br>**(2017) /**<br>**DURATION-**<br>**NEO-2 Trial<sup>78</sup>** | Phase 3 | Adults with T2DM on a<br>stable regimen of metformin | 364 | 54 | 172<br>(47.3) | Exenatide (2.0 mg<br>QW) | Sitagliptin or<br>Placebo | 28 weeks | Change in HbA1c |
| **Knop et al.**<br>**(2023) / OASIS**<br>**1 Trial<sup>55</sup>** | Phase 3 | Obese adults with at least<br>one self-reported dietary<br>weight loss effort | 667 | 50 | 485<br>(72.7) | Semaglutide (50 mg<br>OD) | Placebo | 68 weeks | Percentage change in BW<br>and whether participants<br>reached at least 5% BW<br>reduction |
| **Siskind et al. (2017) / CODEX Trial**<sup>79</sup> | Phase 2 | Adults with a clinical diagnosis of schizophrenia or schizoaffective disorder taking oral clozapine | 28 | - | 10 (35.7) | Exenatide (2 mg QW) | Usual care (Clozapine alone) | 24 weeks | Difference in the proportion of participants with >5% BW reduction |
| **Moretto et al. (2008)**<sup>76</sup> | Phase 2 | Overweight or obese adults with T2DM | 232 | 54 | 102 (44.0) | Exenatide (5 ug or 10 ug BID) | Placebo | 24 weeks | Change in HbA1c |
| **Lingvay et al. (2018)**<sup>60</sup> | Phase 2 | Adults with T2DM and on stable diabetes treatment | 705 | 57 | 326 (46.2) | Semaglutide (0.05, 0.1, 0.2 or 0.3 mg OD) | Liraglutide or Placebo | 26 weeks | Change in HbA1c |
| **Dungan et al. (2016) / AWARD-8 Trial**<sup>64</sup> | Phase 3 | Adults with a BMI of $\leq 45$ kg/m2 with T2DM not optimally controlled with diet and exercise | 300 | 58 | 167 (55.7) | Dulaglutide (1.5 mg QW) | Placebo | 24 weeks | Change in HbA1c |
| **Dejgaard et al. (2019) / Lira Pump Trial**<sup>23</sup> | Phase 3 | Adults with T1DM, treated with CSII, in non-optimal glycemic control | 44 | 50 | 30 (68.2) | Liraglutide (1.8 mg OD) | Placebo | 26 weeks | Change in HbA1c |
| **Gerstein et al. (2019) / REWIND Trial**<sup>65</sup> | Phase 3 | Adults with T2DM whose HbA1c was 9.5% or less, on stable doses of up to two oral glucose-lowering drugs | 9,901 | 66 | 4,589 (46.3) | Dulaglutide (1.5 mg QW) | Placebo | Treatment period is not uniform for all subjects | MACE |
| **O'Neil et al. (2018)**<sup>61</sup> | Phase 2 | Obese adults without diabetes | 957 | 47 | 619 (64.7) | Semaglutide (0.05, 0.1, 0.2, 0.3 or 0.4 mg OD) or Liraglutide (3 mg OD) | Placebo and active control (Liraglutide) | 52 weeks | Percentage weight loss |
| **Lundkvist et al. (2017)**<sup>75</sup> | Phase 2a | Obese adults without diabetes | 50 | 53 | 30 (60.0) | Exenatide (2 mg QW) | Placebo | 52 weeks | Change and percent change in BW |
| **Mosenzon et al. (2019) / PIONEER 5 Trial**<sup>56</sup> | Phase 3a | Adults with T2DM and an eGFR of 30–59 mL/min per 1.73 m2 | 324 | 70 | 168 (51.9) | Semaglutide (14 mg OD) | Placebo | 26 weeks | Change in HbA1c |
| **Wilding et al. (2021)**<sup>63</sup> | Phase 3 | Adults with one or more self-reported unsuccessful dietary efforts to lose weight | 1,961 | 46 | 1,453 (74.1) | Semaglutide (2.4 mg QW) | Placebo | 68 weeks | Percentage change in BW and weight reduction of at least 5% |
| **Lincoff et al. (2023) / SELECT Trial**<sup>4</sup> | Phase 4 | Overweight or obese patients who had preexisting cardiovascular disease but had no history of diabetes | 17,604 | 62 | 12,732 (72.3) | Semaglutide (2.4 mg QW) | Placebo | 136 weeks | Composite of death from cardiovascular causes, nonfatal myocardial infarction, or non-fatal stroke |
| **Husain et al. (2019) / PIONEER 6 Trial**<sup>57</sup> | Phase 3 | Patients with established cardiovascular disease or chronic kidney disease | 3,183 | 66 | 1,007 (31.6) | Semaglutide (14 mg OD) | Placebo | 69 weeks | MACE |
| **Gill et al. (2010)**<sup>72</sup> | Phase 2 | Adults on a stable metformin dose for 30 days or on TZD for 120 days | 54 | 57 | 24 (44.4) | Exenatide (10 µg BID) | Placebo | 12 weeks | Change in mean 24-hour HR |
| **Ghanim et. al (2020)**<sup>32</sup> | Phase 3 | Overweight or obese adults with T1DM, on stable basal and bolus insulin, and had no detectable C-peptide in plasma | 84 | 47 | 60 (71.4) | Liraglutide (1.8 mg OD) | Placebo | 26 weeks | Change in HbA1c |
| **Sorli et al. (2017) / SUSTAIN 1 Trial**<sup>58</sup> | Phase 3a | Adults with T2DM treated with only diet and exercise alone | 387 | 54 | 177 (45.7) | Semaglutide (0.5 mg or 1.0 mg QW) | Placebo | 30 weeks | Change in HbA1c |
| **le Roux et al. (2017)**<sup>40</sup> | Phase 3a | Obese adults with stable bodyweight, with dyslipidemia or hypertension, or both | 2,254 | 48 | 1,714 (76.0) | Liraglutide (3.0 mg OD) | Placebo | 160 weeks | Proportion of individuals with type 2 diabetes at 160 weeks |
| **Davies et al. (2017)**<sup>50</sup> | Phase 2 | Adults with T2DM and insufficient glycemic control | 632 | 57 | 395 (62.5) | Semaglutide (2.5, 5, 10, 20, and 40 mg OD) | Placebo or SC Semaglutide | 26 weeks | Change in HbA1c |
| **Smits et al. (2017)**<sup>47</sup> | Phase 3 | Patients with T2DM | 36 | 61 | 9 (25.0) | Liraglutide (1.8 mg OD) | Placebo | 12 weeks | Change in LF/HF ratio |
| **Idorn et al. (2016)**<sup>33</sup> | Phase 2 | Adults with T2DM and ESRD | 23 | 66 | 8 (34.8) | Liraglutide (0.6 mg OD) | Placebo | 12 weeks | Dose-corrected trough concentration of liraglutide in plasma at the final trial visit |
| **Rubino et al. (2021) / STEP 4 Trial**<sup>53</sup> | Phase 3a | Adults with at least one self-reported unsuccessful dietary effort to lose weight | 803 | 46 | 634 (79.0) | Semaglutide (2.4 mg QW) | Placebo | 68 weeks | Percent change in BW |
| **Grunberger et al. (2012)**<sup>67</sup> | Phase 2 | Adults with T2DM who were anti-hyperglycemic medication-naïve or had discontinued metformin monotherapy | 167 | 57 | 90 (54.9) | Dulaglutide (0.1, 0.5, 1.0 or 1.5 mg QW) | Placebo | 12 weeks | Change in HbA1c |
| **Kim et al. (2013)**<sup>34</sup> | Phase 3 | Adults with prediabetes | 51 | 58 | 33 (64.7) | Liraglutide (1.8 mg OD) | Placebo | 14 weeks | Percentage loss of baseline BW and change in insulin resistance |
| **Kumarathurai et al. (2017)**<sup>37</sup> | Phase 3 | Overweight patients with T2DM and stable CAD | 24 | 63 | 22 (91.7) | Liraglutide (1.8 mg OD) | Placebo | 12 weeks | Change in mean 24-hour SBP and DBP |
| **Kosiborod et al (2023) / STEP HFpEF trial**<sup>3</sup> | | Adults with LVEF ≥ 45%; BMI ≥ 30; NYHA class II, III, or IV symptoms; KCCQ-CSS score $<$ 90 points; 6MWT ≥ 100 m; and at least 1 of the following:<br>elevated LV filling pressures, elevated natriuretic peptide levels plus echocardiographic abnormalities, or hospitalization for HF in the 12 months before screening plus ongoing treatment with diuretics or echocardiographic abnormalities | 529 | 69 | 297 (56.1) | Semaglutide (2.4 mg QW) | Placebo | 52 weeks | Change from baseline in the Kansas City Cardiomyopathy Questionnaire clinical summary score (KCCQ-CSS) |
| **Ferdinand et al. (2014)**<sup>66</sup> | Phase 3 | Patients with T2DM, on ≥1 oral antihyperglycemic medication for ≥1 month | 755 | 57 | 363 (48.1) | Dulaglutide (1.5mg or 0.75mg QW) | Placebo | 26 weeks | Change in mean 24-hour SBP |
| **Kumarathurai et al. (2016)**<sup>36</sup> | Phase 2 | Adults with T2DM and stable CAD | 41 | 62 | 9 (22.0) | Liraglutide (1.8 mg BID) | Placebo | 24 weeks | Change in LVEF |
| **Faber et al. (2015)**<sup>31</sup> |  | Adults with T2DM on monotherapy with metformin or SU or a combination | 20 | 57 | 5 (25.0) | Liraglutide (1.2 mg OD) | Usual diabetic therapy (Metformin or SU or combination) | 10 weeks | Effect on coronary microcirculation, estimated by CFR |
| **Buse et al. (2011)**<sup>70</sup> | Phase 2 | Adults with T2DM, receiving insulin glargine at a minimum of 20 U/day | 259 | 59 | 111 (42.9) | Exenatide (10 μg BID) | Placebo | 30 weeks | Change in HbA1c |
| **Chen et al. (2015)**<sup>83</sup> | Phase 3 | Patients with NSTEMI | 90 | 58 | 24 (26.7) | Liraglutide (1.8 mg OD) | Placebo | 1 week | Change in LVEF |
| **Kumarathurai et al. (2021)**<sup>38</sup> | Phase 2 | Overweight patients with stable CAD, LVEF >40% and T2DM | 30 | 63 | 6 (20.0) | Liraglutide (1.8 mg OD) | Placebo | 24 weeks | Change in diastolic function parameters |
| **Whicher et al (2021)**<sup>49</sup> | Phase 2 | Adults with schizophrenia, schizoaffective or first-episode psychosis who are overweight or obese | 47 | 43 | 13 (27.7) | Liraglutide (3 mg OD) | Placebo | 24 weeks | Estimate of adherence to investigational product |
| **Davies et al. (2015) / SCALE Diabetes Trial**<sup>81</sup> | Phase 3 | Adults with T2DM treated with diet and exercise alone or in combination with 1 to 3 oral hypoglycemic agents | 827 | 55 | 421 (50.9) | Liraglutide (3.0 mg or 1.8 mg OD) | Placebo | 56 weeks | Relative change in BW, proportion of participants losing 5% or more of BW, and proportion losing more than 10% of BW |
| **Kuhadiya et al. (2016)**<sup>35</sup> | Phase 4 | Adults with T1DM, with fasting C-peptide < 0.1 nmol/L | 63 | 43 | 35 (55.6) | Liraglutide (0.6 mg, 1.2 mg or 1.6 mg OD) | Placebo | 12 weeks | Change in mean weekly BG concentrations |
| **de Wit et al. (2014) / ELEGANT Trial**<sup>24</sup> | Phase 2 | Adults with T2DM who had documented weight gain of ≥4% body weight since initiation of insulin treatment. | 50 | 57 | 19 (38.0) | Liraglutide (1.8 mg OD) | Placebo | 26 weeks | Change in BW |
| **Klausen et al. (2022)**<sup>73</sup> | Phase 2 | AUD patients who had a minimum of 5 heavy drinking days in the past 30 days | 127 | 52 | 51 (40.2) | Exenatide (2 mg QW) | Placebo | 26 weeks | Reduction in number of heavy drinking days. |
| **Mensberg et al. (2016)**<sup>44</sup> | Phase 2 | Adults with T2DM treated with diet and/or metformin and sedentary lifestyle | 33 | 57 | 10 (30.3) | Liraglutide (1.8 mg OD) | Placebo | 16 weeks | Change in HbA1c |
| **Miyagawa et al. (2015)**<sup>68</sup> | Phase 3 | Adults who were OAM-naïve or had discontinued OAM monotherapy | 492 | 57 | 91 (18.5) | Liraglutide (0.9 mg OD) | Placebo or Dulaglutide | 26 weeks | Change in HbA1c |
| **Lundgren et al. (2021)**<sup>42</sup> | Phase 3 | Obese adults | 195 | 43 | 124 (63.6) | Liraglutide (3 mg OD) | Placebo | 48 weeks | Change in BW |
| **Guja et al. (2017) / DURATION-7 Trial**<sup>77</sup> | Phase 3 | Adults with T2DM, on an insulin regimen in combination with diet and exercise or with metformin | 461 | 58 | 240 (52.1) | Exenatide (2 mg QW) | Placebo | 28 weeks | Change in HbA1c |
| **Matikainen et al. (2018)**<sup>43</sup> | Phase 2 | Obese patients with T2DM, under treatment with lifestyle changes or metformin | 22 | 62 | 6 (27.3) | Liraglutide (1.8 mg OD) | Placebo | 16 weeks | Changes in weight, HbA1c, β-hydroxybutyrate and SBP |
| **Liakos et al. (2018)**<sup>41</sup> | Phase 2 | Adult patients with inadequately controlled T2DM despite stable antihyperglycemic therapy | 62 | 61 | 21 (33.9) | Liraglutide (1.2 mg OD) | Placebo | 5 weeks | Change in 24-hour ambulatory SBP |
| **Pi-Sunyer et al. (2015)**<sup>45</sup> | Phase 3 | Obese adults who had stable body weight | 3,731 | 45 | 2,928 (78.5) | Liraglutide (3 mg OD) | Placebo | 56 weeks | Weight change, the proportion of patients who lost at least 5%, and those who lost more than 10% of their baseline BW |
| **Larsen et al. (2017)**<sup>39</sup> | Phase 2 | Overweight or obese adults with a schizophrenia spectrum disorder who were receiving clozapine or olanzapine and had prediabetes | 103 | 42 | 37 (38.1) | Liraglutide (1.8 mg OD) | Placebo | 16 weeks | Change in glucose tolerance |
| **Liutkus et al. (2010)**<sup>74</sup> | Phase 3 | Adults with T2DM, with suboptimal glycemic control while taking stable doses of TZD therapy or combined metformin and TZD therapy | 165 | 55 | 67 (40.6) | Exenatide (10 mcg BID) | Placebo | 26 weeks | Change in HbA1c |
| **Bojer et al. (2021)**<sup>29</sup> | Phase 3 | Adults with T2DM with signs of diastolic dysfunction | 40 | 62 | 8 (20.0) | Liraglutide (1.8 mg OD) | Placebo | 18 weeks | Change in left ventricle ePFR and left atrium PEF |
| **Sandsdal et al. (2023)**<sup>46</sup> | Phase 3 | Obese adults | 195 | 43 | 124 (63.6) | Liraglutide (3 mg OD) | Placebo or Exercise | 52 weeks | Change in MetS-Z, abdominal obesity, and CRP |
| **Wadden et al. (2019)**<sup>48</sup> | Phase 2 | Obese adults with prior lifetime weight loss effort with diet and exercise | 150 | 48 | 119 (79.3) | Liraglutide (3 mg OD) | Behavioral Therapy | 52 weeks | Change in BW |
| **Garvey et al (2022) / STEP 5 Trial**<sup>54</sup> | Phase 3 | Obese adults with at least one self-reported unsuccessful dietary effort to lose body weight | 304 | 47 | 236 (77.6) | Semaglutide (2.4 mg QW) | Placebo | 104 weeks | Percentage change in BW and achievement of weight loss of at least 5% |
| **Rosenstock et al. (2018) / FREEDOM-1 Trial**<sup>80</sup> | Phase 3 | Adults with T2DM receiving stable treatment with diet and exercise alone or with metformin, SUs, or pioglitazone | 460 | 56 | 188 (40.9) | Exenatide (40 µg or 60 µg OD) | Placebo | 39 weeks | Change in HbA1c |
| **Mok et al. (2023) / BARI-OPTIMISE Trial**<sup>25</sup> | Phase 3a | Patients with poor weight loss after metabolic surgery and a suboptimal nutrient-stimulated GLP-1 response | 70 | 46 | 52 (74.3) | Liraglutide (3.0 mg OD) | Placebo | 24 weeks | Percentage change in BW |
| **Rodbard et al. (2018) / SUSTAIN 5** | Phase 2 | Adults with T2DM receiving stable basal insulin therapy alone or in | 396 | 47 | 174 (43.9) | Semaglutide (0.5 mg or 1.0 mg QW) | Placebo | 30 weeks | Change in HbA1c |

| Trial<sup>59</sup> |  | combination with metformin |  |  |  |  |  |  |  |
| --- | --- | --- | --- | --- | --- | --- | --- | --- | --- |
| Lind et al. (2015) / MDI Liraglutide trial<sup>26</sup> | Phase 3a | Overweight or obese adults with T2DM treated with multiple daily insulin injections | 124 | 59 | 44 (35.5) | Liraglutide (1.8 mg OD) | Placebo | 24 weeks | Change in HbA1c |
| Gullaksen et al. (2023)<sup>62</sup> | Phase 3 | Patients with T2DM, with established CVD, heart failure or CKD | 80 | 64 | 19 (23.8) | Semaglutide (1.0 mg QW) | Placebo or Empagliflozin | 16 weeks | Improvement in kidney tissue oxygenation |
| Dushay et al. (2012)<sup>71</sup> | Phase 3 | Overweight or obese adult women without diabetes | 41 | 70 | 41 (100.0) | Exenatide (10 ug BID) | Placebo | 16 weeks | Change in body weight and BMI |
| Astrup et al. (2009)<sup>28</sup> | Phase 3 | Obese adults | 564 | 48 | 429 (76.1) | Liraglutide (1.2 mg, 1.8 mg, 2.4 mg or 3 mg OD) | Placebo or Orlistat | 20 weeks | Change in BW |
| Armstrong et al. (2006) / LEAN Trial<sup>27</sup> | Phase 3 | Overweight patients who have clinical evidence of non-alcoholic steatohepatitis | 52 | 46 | 31 (59.6) | Liraglutide (1.8 mg OD) | Placebo | 48 weeks | Number of patients with paired liver biopsies and patients with resolution of non-alcoholic steatohepatitis |
| Apovian et al. (2010)<sup>69</sup> | Phase 2 | Overweight or obese adults with T2DM on a stable dose of metformin or SU, and with a history of stable body weight | 194 | 50 | 121 (62.4) | Exenatide (10 ug BID) | Placebo and Lifestyle Modifications | 24 weeks | Change in BW |

**Abbreviations:** AUD=alcohol use disorder; BG=blood glucose; BID=twice a day; BMI=body mass index; BW=body weight; CAD=coronary artery disease; CFR=coronary flow reserve; CHF=congestive heart failure; CKD=chronic kidney disease; CRP=C-reactive protein; CSII=continuous subcutaneous insulin infusion; CVD=cardiovascular disease; DBP=diastolic blood pressure; ePFR=early peak filling rate; ESRD=end stage renal disease; HR=heart rate; LF/HF=low frequency to high frequency; LVEF=left ventricular ejection fraction; MACE=major adverse cardiovascular events; NSTEMI=non-ST elevation myocardial infarction; OAM=oral antihyperglycemic medication; OD=once a day; PEF=passive emptying fraction; QW=once a week; SBP=systolic blood pressure; SC=subcutaneous; SU=sulfonylurea; T1DM=type 1 diabetes mellitus; T2DM=type 2 diabetes mellitus; TZD=thiazolidinedione

Liraglutide had the most RCTs with 30 studies^20–49^ having it as the intervention. There were 16 RCTs^3, 4, 50–63^ done for Semaglutide and 5 for dulaglutide.^64–68^ Exenatide had 12 RCTs.^69–80^ Thirty-eight (38) ^RCTs21, 24, 26, 29, 31, 33, 36–38, 41, 43, 44, 47, 51, 55–60, 62, 64–70, 72, 74, 76–78, 80, 81^ were done exclusively among patients with DM. Among these RCTs, majority^21, 24, 26, 41, 43, 44, 47, 51, 55, 60, 64, 68,74, 76–78, 80–82^ enrolled exclusively T2DM patients. Seven (7) RCTs^20, 25, 27, 49, 50, 79, 83^ enrolled both patients with or without diabetes. The remaining eighteen RCTs did not include patients with DM, three (3)^34, 39, 40^ of which included exclusively patients with prediabetes.

Among the RCTs in this study, 61 have data for SBP^3, 4, 20, 21, 23–29, 31–46, 48–51, 53–81, 83^. Conversely, 55 RCTs have data for DBP. The method of BP measurement varied among the RCTs. Five (5) RCTs^29, 37, 41, 66, 72^ used continuous or 24-hour ambulatory blood pressure monitoring (ABPM). Among these, Bojer et al. (2021)^29^ performed orthostatic BP measurement with supine and standing BP readings. The remaining 58 RCTs did not employ continuous BP monitoring, but instead utilized routine BP measurements, which were performed at specified intervals; methodology varied among these studies: 22 RCTs^21, 28, 31, 34, 35, 38, 44, 46, 47, 50, 51, 53–55, 57, 61, 64, 65, 68, 75, 78, 79^ used seated blood pressure, Gullaksen et al. (2023)^62^ used supine BP, Wadden et al. (2019)^48^ used standing BP, Dushay et al. (2012)^71^ did not mention the posture of the patient but used a Dynamap automated monitoring device, and 33 other RCTs did not explicitly mention the method of routine BP measurement.

Forty-four (44) RCTs included patients with CKD, but many trials employed a varying cutoff eGFR to exclude patients with more severe CKD: Klausen et al., 2022^73^ excluded patients with eGFR = 50 mL/min/1.73 m^2^ and/or microalbuminuria. Gullaksen et al., 2023^62^ excluded patients with eGFR < 45 mL/min/1.73 m^2^. STEP 6^50^ excluded patients with eGFR < 30 mL/min/1.73 m^2^ if the patient has T2DM (only in Japan) but excluded patients with eGFR < 15 mL/min/1.73 m^2^ if the patient has no T2DM. Eighteen (18) RCTs^20, 21, 25, 41, 42, 46, 49, 51, 55–58, 64–66, 77–79^ excluded patients with eGFR < 30 mL/min/1.73 m^2^; among these 18 trials, PIONEER 5^55^ specifically excluded patients with rapidly progressive renal disease or known nephrotic albuminuria (> 2200 mg/24 hr or > 2200 mg/g), Ferdinand et al., 2014^66^ also excluded patients with eGFR = 30 mL/min/1.73 m^2^, DURATION-7^77^ further excluded patients on metformin with eGFR < 60 mL/min/1.73 m2, serum creatinine >/= 1.5 mg/dL in men, or >/= 1.4 mg/dL in women. Six (6) RCTs^3, 4, 52–54, 63^ excluded patients with eGFR < 15 mL/min/1.73 m^2^. Ten (10) RCTs did not mention CKD or renal disease as an exclusion criterion. Idorn et al. (2016)^33^ specifically included patients with end-stage renal disease. Six (6) RCTs^24, 26, 27, 35, 39, 67^ used exclusively a serum creatinine (sCr) level as a cutoff to exclude patients with CKD, with three (3) RCTs^26, 27, 39^ excluding patients with sCr > 150 μmol/L; of these 3, Larsen et al., 2017^39^ excluded patients with macroalbuminuria. The 3 other RCTs have sCr cutoff values as follows: ELEGANT^24^ excluded patients with sCr > 130 μmol/L, Kuhadiya et al. (2016)^35^ excluded patients with sCr > 1.5 mg/dL, and Grunberger et al. (2012)^67^ excluded patients wtih sCr >/= 1.5 mg/dL in men or >/= 1.4 mg/dL in women. However, using a serum creatinine value as an exclusion criterion can end up including patients with CKD despite having a normal sCr, which tends to occur in the elderly.^84^

Eighteen (18) RCTs excluded patients with CKD, twelve (12)^23, 29, 36–38, 43, 47, 59, 60, 75, 80, 83^ of which used an eGFR cutoff of < 60 mL/min/1.73 m^2^, while six (6)^31, 34, 44, 48, 70, 76^ did not use an eGFR cutoff but excluded patients with known CKD. For the remaining trial, which is Astrup et al., 2009^28^, it was not clear if CKD patients are excluded since “major medical conditions” was an exclusion criterion, but it did not explicitly mention as to whether CKD is a major medical condition.

The reporting of baseline blood pressure varied among RCTs. Among them, nine (9)^36, 38, 46, 52, 54, 59, 60, 71, 78^ RCTs reported a mean baseline SBP that included all treatment groups. Of these 9 RCTs, five (5)^46, 52, 54, 71, 78^ have a mean SBP of 120-129 mmHg at baseline, three (3)^36, 59, 60^ have a mean baseline SBP 130-139, and only one^38^ have mean baseline SBP >/= 140. Forty-five (45) studies did not report a mean across all treatment groups but reported means of each treatment arm. Of these 45 RCTs, two (2) have treatment arms with mean SBPs belonging to different ranges. Conversely, the other 43 RCTs have either only 1 treatment arm or have multiple treatment arms with mean SBPs belonging to the same range: 18 RCTs^20, 21, 23, 28, 32, 35, 39, 40, 42, 45, 53, 63, 66, 67, 69, 79, 83^ have a treatment arm with mean SBP of 120-129 mmHg at baseline, 24 RCTs^3, 4, 24, 25, 27, 29, 37, 43, 44, 47–51, 56, 62, 64, 65, 70, 72, 74, 75, 77, 80^ have a treatment arm with mean baseline SBP 130-139, and only one^31^ have a treatment arm with mean baseline SBP >/= 140. The remaining nine (9)^26, 33, 41, 55, 57, 58, 61, 68, 73^ RCTs did not mention any baseline SBP.

### Systolic blood pressure reduction

#### Semaglutide

Semaglutide significantly reduced SBP with a MD of −3.40, (95% CI, −4.22 to −2.59), p<0.001, (I^2^ 87.78; p<0.001). Subgroup analysis based on the mode of administration showed significant SBP reduction for both oral (MD: −4.06, 95% CI (−5.19 to −2.93), p<0.001) and subcutaneous (MD: −3.40, 95% CI (−4.13 to −2.31), p<0.001) semaglutide. **(See figure 1)** Subgroup analysis based on treatment duration showed significant SBP reduction for patients who received treatment for more than 26 weeks (MD: −4.23, 95% CI (−5.02 to −3.44), p<0.001) compared to those who received the treatment for 26 weeks or less (MD: −0.65, 95% CI (−1.13 to −0.16), p<0.001), p-interaction<0.001). **(See figure 2)** Subgroup analysis for subcutaneous Semaglutide based on dosing showed significant SBP reduction for patients who received a dose of >1 mg (MD: −4.57, 95% CI (−5.89 to −3.26), p<0.001) compared to those who received a dose of 1 mg or less (MD: −2.52, 95% CI (−3.44 to −1.59), p<0.001), (p-interaction NS). **(See figure S3)** Subgroup analysis based baseline mean SBP showed significant SBP reduction for patients who had a mean SBP between 120-129 mmHg (MD: −5.19, 95% CI (−7.28 to −3.09), p<0.001) compared to those who had a mean SBP between 130-139 mmHg (MD: −2.66, 95% CI (−3.84 to − 1.48), p<0.001), (p-interaction<0.04). **(See figure 3)** Subgroup analysis based on DM status showed significant reduction in SBP among those without DM (MD: −4.60, 95% CI −5.61 to - 3.59, p<0.01) versus among those with DM (MD: −2.15, 95% CI −3.14 to −1.15, p<0.01), (p-interaction <0.01). **(See figure 4)**

#### Liraglutide

Liraglutide significantly reduced SBP with a MD of −2.61 (95% CI, −3.48 to −1.74), p<0.001. Subgroup analysis based on treatment duration showed significant SBP reduction for both those who received treatment for more than 24 weeks (MD: −2.35, 95% CI (−3.44 to −1.25), p<0.001) and to those who received the treatment for 24 weeks or less (MD: −3.11, 95% CI (−4.40 to − 1.83), p<0.001), p-interaction<NS). **(See figure 5)** Subgroup analysis based on dosing showed similar significant SBP reduction for patients who received a dose of >1.8 mg (MD: −2.65, 95% CI (−4.40 to −0.89), p<0.001) and to those who received a dose of 1.8 mg or less (MD: −2.68, 95% CI (−3.46 to −1.91), p<0.001), p-interaction NS). **(See figure S4)** Subgroup analysis based on the mean baseline SBP showed significant SBP reduction for patients who had baseline SBP of 120-129 mmHg (MD: −2.64, 95% CI (−3.36 to −1.93), p<0.001) than those who had a mean baseline SBP of 130-139 mmHg (MD: −0.98, 95% CI (−3.03 to 1.07), p=0.035). **(See figure S5)** Subgroup analysis based on DM status showed significant reduction in SBP for both those without DM (MD: −2.92 mmHg, 95% CI −4.54 to −1.30, p<0.01) versus among those with DM (MD: −2.57, 95% CI −3.39 to −1.76, p<0.01), (p-interaction NS). **(See figure S6)**

#### Dulaglutide

Dulaglutide significantly reduced systolic blood pressure with a MD of −1.46 (95% CI, −2.20 to − 0.72), p<0.001. Subgroup analysis based on treatment duration both showed significant SBP reduction for patients who received treatment for 24 weeks or more (MD: −1.14, 95% CI (−2.09 to −0.19), p<0.001) compared to those who received the treatment for less than 24 weeks (MD: - 2.00 95% CI (−2.20 to −0.72), p<0.001), p-interaction NS). **(See figure 6)** Subgroup analysis based on dosing showed significant SBP reduction for patients who received a dose of 1 mg or more (MD: −1.90, 95% CI (−2.92 to −0.88), p<0.001) compared to those who received a dose of less than 1 mg (MD: −0.84, 95% CI (−1.81 to −0.13), p<0.001), p-interaction NS). **(See figure S7)** Subgroup analysis based on the mean baseline SBP showed significant SBP reduction for patients who had baseline SBP of 120-129 mmHg (MD: −2.11, 95% CI (−2.79 to −1.43), p<0.001) than those who had a mean baseline SBP of 130-139 mmHg (MD: −1.11, 95% CI (−2.28 to 0.05), p=0.06). **(See figure S8)**

#### Exenatide

Exenatide significantly reduced SBP with a MD of −3.36, (95% CI, −3.63 to −3.10), p<0.001. Subgroup analysis based on treatment duration both showed significant SBP reduction for patients who received treatment for 24 weeks or more (MD: −3.37, 95% CI (−3.63 to −3.11), p<0.001) compared to those who received the treatment for less than 24 weeks (MD: −1.63, 95% CI (−5.63 to 2.36), p=0.42), p-interaction NS). **(See figure S9)** Subgroup analysis based on the mean baseline SBP showed similar significant SBP reduction for patients who had baseline SBP of 120-129 mmHg (MD: −2.75, (95% CI −5.22 to −0.27, p=0.03) and those who had a mean baseline SBP of 130-139 mmHg (MD: −3.37, 95% CI (−3.73 to −3.00), p=0.00). (**See figure 10)**

### Diastolic blood pressure reduction

Semaglutide (MD: −0.72, (95% CI, −1.54 to −0.10, p=0.08), liraglutide (MD: −0.23 (95% CI, −0.59 to 0.12, p=0.68), and dulaglutide (MD: 0.27 (95% CI, −0.05 to 0.59, p=0.10) failed to reduce DBP. Exenatide modestly reduced DBP (MD: −0.94 (95% CI, −1.78 to −0.10, p=0.03.

#### Meta-regression analysis

Meta-regression analysis revealed that mean change in weight (p=0.02, tau^2^ 3.047, R^2^ analog=37.65), **(Figure S11)** mean change in A1c (p=0.010, tau^2^ 2.989, R^2^ analog=28.27) **(Figure S12)** and mean change in BMI (p=0.00, tau^2^ 0.165, R^2^ analog=92.57) **(Figure S13)** were associated with the treatment effect for SBP reduction for semaglutide. For exenatide, only the mean change in A1c affected the treatment effect (p=0.03, tau^2^ 9.8, R^2^ analog=80.20). For dulaglutide, mean change in A1c (p=0.002, tau^2^ 0.1308, R^2^ analog=81.80) and mean change in weight (p=0.00, tau^2^ 6.6, R^2^ analog=100.00) affected the treatment effect. These factors did not affect the treatment effect for liraglutide.

## DISCUSSION

We completed a detailed meta-analysis and meta-regression on the efficacy of the four common GLP-1RAs in reducing blood pressure. We established that GLP-1RAs reduced SBP by 1.46 to 3.40 mmHg. Semaglutide, liraglutide, dulaglutide and exenatide all reduced SBP to a certain degree. Through regression analyses, we established that mean change in weight, mean change in A1c, and mean change in BMI were directly associated with SBP reduction. Subgroup analyses done between dose and route of semaglutide and its effect on SBP revealed statistically significant, but clinically modest SBP reduction that was comparable between two dose groups of semaglutide (high vs low dose regimen) and between two different routes of administration (oral vs subcutaneous). Those with lower mean baseline SBP experienced greater BP lowering effect. Furthermore, greater SBP reduction was noted among patients who received longer duration of semaglutide treatment compared to those who had shorter treatment duration. Comparing patients with vs without T2DM, we found that semaglutide modestly reduced SBP with statistical significance among those without T2DM compared to those with T2DM, while liraglutide modestly reduced SBP in both cohorts. Contrary to its promising effects in SBP lowering, GLP-1RA has very minimal to no effect on DBP. Of the four GLP-1RAs, only exenatide reduced DBP which was more pronounced to those without T2DM.

### SBP reduction, Semaglutide

We established that semaglutide demonstrated a statistically significant but clinically modest reduction in SBP compared to placebo by 3.40 mmHg. This has been demonstrated by multiple large RCTs that enrolled > 1,000 participants such as PIONEER 6^56^, SELECT^4^, and Wilding et al. (2021)^63^, as well as other RCTs that enrolled hundreds such as STEP 2^51^, STEP 6^50^, OASIS 1^54^, and O’Neil et al. (2018)^61^. There was substantial heterogeneity in our results (I2 = 89.18%), and this can be explained by some RCTs in our study, such as STEP 5^53^, STEP HFpEF^3^, and SUSTAIN 1^57^, which did not demonstrate a statistically significant reduction in SBP. The results of our study were consistent with a recent meta-analysis done by Kennedy et al. (2023)^85^, which included 6 RCTs with 4,744 participants; this meta-analysis showed that semaglutide decreased SBP by 4.83 mmHg (95% CI, −5.65 to −4.02).

The route of administration of semaglutide had no significant effect on SBP reduction, as our study showed that orally administered semaglutide reduced SBP by 4.06 mmHg whereas subcutaneous administration reduced SBP by 3.22 mmHg (p interaction NS). This is consistent with an RCT done by Davies et al. (2017)^59^, which compared subcutaneous semaglutide 1 mg to varying doses of oral semaglutide (2.5 to 40 mg); a similar reduction in SBP was observed between the two routes: 5.7 mmHg with subcutaneous semaglutide vs 5.4 to 7.8 mmHg among different oral semaglutide doses. Notedly, oral semaglutide differed from subcutaneous administration in terms of the dosage used and frequency of administration: oral semaglutide is given daily with a higher dose, whereas subcutaneous semaglutide is administered weekly.

Prolonged administration of semaglutide had a greater impact on SBP, thereby suggesting a duration-dependent effect: there was a reduction of SBP by 4.25 mmHg among trials that administered semaglutide for > 26 weeks, compared to trials that administered semaglutide for </= 26 weeks, with SBP reduction of 0.55 mmHg. Four RCTs that administered semaglutide for less than 26 weeks had no significant reduction in SBP.^55, 59, 60, 62^ On the other hand, SBP reduction among RCTs that administered semaglutide for > 26 weeks varied, as some trials^3, 53, 57^ failed to demonstrate significant SBP reduction but most other trials with duration > 26 weeks^4, 50, 51, 54, 56, 61, 63^ were able to demonstrate this treatment effect.

There was a marked reduction in SBP with higher doses of subcutaneous (SC) semaglutide compared to lower doses. O’Neil et al.^61^ successfully demonstrated a significant SBP reduction (mean −4.49 to −2.88 mmHg) between higher versus lower doses. Another trial, SUSTAIN 5^58^, showed that semaglutide 0.5 mg did not significantly reduce SBP by 3.30 mmHg (95% CI −6.91 to 0.31), but semaglutide 1 mg showed a significant reduction in SBP by 6.30 mmHg (95% CI −9.92 to −2.68), suggesting a possible dose-dependent effect on SBP reduction between 0.5 mg and 1 mg. Notably, patients with lower baseline SBP had a trend towards more marked SBP reduction than those with higher baseline SBP, with mean SBP reduction 5.19 mmHg (95% CI −7.28 to −3.09) among studies with mean SBP 120-129 mmHg vs studies with mean SBP 130-139 mmHg, which showed mean SBP reduction 2.66 (95% CI −3.84 to −1.48). The effect was driven mainly by 2 RCTs with mean SBP 120-129 mmHg, OASIS 1^54^ and Wilding et al., (2021)^63^, which had a mean SBP reduction of 6.30 and 5.94 mmHg, respectively.

### SBP reduction, Liraglutide

Liraglutide demonstrated a statistically significant, but clinically modest SBP reduction compared to placebo with a MD of −2.61 mmHg. Two large RCTs that recruited > 1,000 participants, such as le Roux et al., 2017^40^ and Pi-Sunyer et al., 2015^45^ were able to successfully demonstrate this modest reduction in SBP. This effect was consistent with the finding of a pooled analysis of 6 RCTs, which enrolled 2,783 patients with type 2 diabetes mellitus, as there was a mean SBP reduction of 2.7 and 2.9 mmHg among patients with liraglutide 1.2 mg and 1.8 mg, respectively.^86^ Another meta-analysis that included 18 RCTs with 7,616 individuals was also consistent with our study’s results, as there was mean SBP reduction of 3.18 mmHg with liraglutide treatment (95% CI −4.32 to −2.05) compared to placebo.^14^ Unlike semaglutide, treatment duration did not have a significant effect on SBP reduction. This suggests that liraglutide may only need to be given for at least 5 weeks for it to exert a decrease in SBP, as seen in Liakos et al., 2018^41^. Varying doses of liraglutide decreased SBP modestly with statistical significance compared to placebo, but their effects were not significantly different compared to each other. This suggests that modest SBP reduction may be attained by using a dose as low as 0.6 mg, as Kuhadiya et al. (2016)^35^ has shown this statistically significant but clinically modest treatment effect on SBP using this dose, with decrease of SBP by 2.50 mmHg (95% CI −4.97 to −0.03).

A lower baseline SBP showed an impact on SBP reduction by liraglutide, as RCTs with mean baseline SBP 120-129 mmHg showed a statistically significant but clinically modest decrease in SBP of 2.64 (95% CI −3.36 to −1.93), whereas RCTs with mean baseline SBP 130-139 mmHg showed a trend towards, albeit nonsignificant decrease in SBP of 0.98 mmHg (95% CI −3.03 to 1.07). This was in contrast to a correlation analysis of an RCT of liraglutide for patients with type 2 diabetes, as it showed that higher baseline SBP was associated with a greater SBP reduction with liraglutide treatment; this study attributed the finding to non-systematic measurement errors, in which a single baseline BP measurement varies from the patient’s long-term average BP, a phenomenon known as “regression to the mean”.^87^ Further studies may need to be conducted to determine the relationship of baseline SBP and SBP reduction on liraglutide.

### SBP reduction, Dulaglutide

Dulaglutide modestly reduced SBP with a MD of −1.46 mmHg. This effect was driven by REWIND^82^ and Ferdinand et al., (2014)^66^ trials; however, some RCTs such as AWARD-8^64^ and Miyagawa et al., 2015^68^, did not show statistically significant decrease in SBP. The results of our study were consistent with a meta-analysis of 6 RCTs on dulaglutide, which showed an SBP reduction of 2.6 mmHg (95% CI −3.8 to −1.5) compared to placebo.^88^ Treatment duration of dulaglutide did not significantly alter the magnitude of SBP reduction. Dulaglutide dose was shown to have an effect on SBP reduction. Studies that used dulaglutide dose of less than 1 mg had a trend towards, but nonsignificant, reduction of SBP by 0.84, whereas RCTs that used dulaglutide dose of 1 mg and above had a modest decrease in SBP of 1.90 mmHg (95% CI −2.92 to −0.88). This can be explained by Miyagawa et al., 2015^68^, which utilized a dose of 0.75 mg but had a nonsignificant increase in SBP 0.09 mmHg (95% CI −0.12 to 0.30). These findings are suggestive of a dose-dependent response of dulaglutide and SBP.

### SBP reduction, Exenatide

Exenatide modestly reduced SBP with a MD of 3.36 mmHg. Our results were consistent with an earlier meta-analysis by Wang et al. (2013),^89^ which included 7 RCTs that compared exenatide with placebo among 1,334 patients with T2DM, with a statistically significant but clinically modest SBP reduction of 5.24 mmHg (95% CI −6.88 to −3.59). However, an RCT that used dapagliflozin and exenatide demonstrated no SBP reduction among patients treated with SBP alone, but patients treated with dapagliflozin and empagliflozin had the greatest decrease in SBP of 6.4 mmHg and 6.7 mmHg (after 1.5 and 16 weeks respectively), compared to patients treated with empagliflozin alone, as they had a decrease in SBP of 4.5 mmHg and 1.8 mmHg (after 1.5 and 16 weeks, respectively); these results indicate a synergistic effect on SBP reduction by an SGLT2i and a GLP-1RA with multiple mechanisms such as non-osmotic sodium storage, changes in baroreceptor reflex setpoint or renal sympathetic nerve activity, or a central neural pathway.^90^

Modest SBP reduction was noted among RCTs with longer treatment duration of at least 24 weeks. Our results indicate a duration-dependent effect of exenatide, in which SBP reduction may only occur if administered for at least 24 weeks.

### DBP reduction

Of the GLP-RAs included in our meta-analysis, we found that only exenatide reduced DBP with statistical significance, albeit clinically modest with a mean difference of −0.94 mmHg (95% CI, −1.78 to −0.10), p=0.03), driven mainly by the Moretto et al. (2008) and Buse et al. (2011) trials. These findings are supported by previous meta-analyses, which also found a reduction in blood pressure with exenatide only.^13, 91^ However, a meta-analysis by Hu et al. (2020)^92^ found a statistically significant decrease in DBP with GLP-RAs compared to baseline, although it must be noted that this meta-analysis additionally included albiglutide and taspoglutide, which are not included in our study, and also included active comparators such as insulin, sulfonylureas, and DPP-4 inhibitors. Another meta-analysis by Wang et al. (2013)^89^ also found a statistically significant decrease in DBP with liraglutide in contrast to our study, but also found the same treatment effect of DBP reduction as our study with exenatide.

Our meta-analysis and previous studies demonstrate that systolic BP, not diastolic BP, is the predominant component GLP-1 receptor-mediated BP control, mainly due to its effects on extracellular volume homeostasis.^93^ Although GLP-1 receptor agonists exert vasodilatory effects through production of NO, GLP-1RAs have also been found to improve arterial stiffness, which may attenuate the BP-lowering effect of GLP-1RAs on diastolic blood pressure.^94, 95^

Weight loss is a notable effect of GLP-1RAs, as shown in multiple RCTs and meta-analyses^96, 97^ and weight loss decreases SBP by 5 mmHg among patients with hypertension and 2-3 mmHg among patients with normotension, with a dose-response relationship of around 1 mmHg per kg of weight loss.^98^ This effect of weight loss on SBP reduction was demonstrated in our meta-regression analysis, which revealed that mean weight change was associated with modest SBP reduction for RCTs that utilized semaglutide (p=0.02, tau2 3.047, R2 analog=37.65) and dulaglutide (p=0.00, tau2 6.6, R2 analog=100.00). However, it was not demonstrated in studies that used exenatide and liraglutide. The shorter half-lives of these two medications could potentially explain the lack of association, especially that exenatide and liraglutide are administered daily and dulaglutide and subcutaneous semaglutide are administered weekly^99^, but more studies may be needed to further elucidate the effect of weight reduction on SBP reduction. The aforementioned meta-analysis by Hu et al. (2020)^92^, showed that weight loss was associated with net change in SBP and even in placebo-corrected SBP changes. A meta-analysis that included 6 RCTs on dulaglutide demonstrated that SBP reduction was mostly mediated by the weight-independent effects of dulaglutide, as only 36% of SBP reduction was mediated by weight loss^88^, thus indicating that dulaglutide decreases SBP not just through its effect on weight but also through other mechanisms.

Our study revealed that mean change in BMI was associated with the treatment effect for SBP reduction for semaglutide only (p=0.00, tau2 0.165, R2 analog=92.57) but not for other GLP-1RAs. Interestingly, while weight loss was associated with SBP reduction for dulaglutide, it was not consistent with BMI reduction as there was no association with SBP reduction, even though weight is a component of BMI. This could be explained by the fact that BMI (expressed in kg/m2) is also determined by height, and that BMI is inversely proportional to the square of height.^105^ The effect of height could possibly blunt the effect of BMI on SBP reduction, hence the lack of association with dulaglutide, but more studies may need to be done to determine the effect of BMI on SBP reduction.

Meta-regression analysis revealed that mean change in A1C was associated with the treatment effect for SBP reduction for semaglutide (p=0.010, tau2 2.989, R2 analog=28.27), exenatide (p=0.03, tau2 9.8, R2 analog=80.20), and dulaglutide (p=0.002, tau2 0.1308, R2 analog=81.80), but not for liraglutide. The previously mentioned meta-analysis by Hu et al. (2020)^92^ showed that HbA1c change was associated with net change in SBP and even in placebo-corrected SBP changes. However, after performing adjustment for weight change, the association between A1C reduction and SBP reduction became insignificant; this suggests that the effect of A1C reduction on BP changes may depend on weight reduction, considering that glycemic control can be achieved better with weight loss.^92^

### Strength and limitations

To our knowledge, this is the first meta-analysis and meta-regression to comprehensively report on the efficacy of GLP-1RAs in reducing blood pressure. There are several limitations that are important to note. This is a study-level meta-analysis, and we could not access individual patient data. Additional limitations include heterogeneity in GLP-1RA studies. Publication bias may also be present, the extent of which could not fully be quantified. However, every effort possible was made to limit bias by utilizing a robust analytical approach to adjust for potential moderators through subgroup analyses and regression analysis.

### Conclusions

GLP-1RAs reduced SBP from 1.46 to 3.40 mmHg. DBP reduction from GLP-RAs was very minimal to none. Mean changes in weight, BMI and A1c directly drive the reduction in SBP. SBP reduction for semaglutide was more marked for patients on prolonged therapy, those with lower baseline mean SBP and for patients without DM. SBP reduction of liraglutide and dulaglutide were more pronounced among patients with a lower baseline mean SBP. Only exenatide reduced DBP with statistical significance, albeit the treatment effect was clinically modest. Further study is needed to elucidate the potential mechanisms of this blood pressure effects from GLP-1RA.

### Perspectives

GLP-1RAs, including semaglutide, liraglutide, exenatide, and dulaglutide, exhibit statistically significant but clinically modest efficacy in reducing SBP independently of glycemic control. Semaglutide leads with the greatest SBP reduction, influenced by factors such as weight, baseline SBP, and diabetes status. Liraglutide and dulaglutide also effectively lower SBP, while exenatide additionally impacts DBP. These findings suggest that GLP-1RAs present a promising approach for cardiovascular health management, warranting further investigation into their precise mechanisms. Their weight-independent effects, potential for personalized therapy, and emerging mechanistic insights pave the way for broader application in diverse patient populations, ultimately improving cardiovascular health across a wider range.

### Novelty and relevance

#### What is new?

- SBP reduction from the use of GLP-1RAs is significant, independent of its effects on glycemia.
- Semaglutide demonstrated superior SBP lowering with greater change in HbA1c and body mass index compared placebo but with less SBP lowering among patients with diabetes.
- DBP reduction was not observed among the GLP-1RA with the exception of a marginal decrease in DBP in patients taking exenatide.

#### What is relevant?

The class of GLP-1RA provide modest improvement in systolic blood pressure and may serve as a useful strategy for patients with overweight/obesity and hypertension.

### Clinical/Pathophysiological implications?

- Mechanisms resulting in the BP lowering effect remain unknown, and the clinical benefit resulting from BP reduction specifically from GLP-1RA require further investigation.
- Future research directions encompass investigating GLP-1RA-induced SBP reduction mechanisms through mechanistic studies, gene expression analysis, and advanced imaging; examining the impact of patient characteristics on response and conducting trials targeting specific subpopulations; assessing long-term effects and clinical outcomes through observational studies and RCTs; exploring novel GLP-1RA formulations and targets, including extended half-lives and additional receptors; and leveraging emerging technologies for personalized treatment strategies by integrating data from diverse sources.
- Addressing these areas promises a deeper understanding of GLP-1RAs in blood pressure control, enhancing cardiovascular outcomes in T2DM and associated conditions

## Abbreviations

BP: blood pressure
BMI: body mass index
DBP: diastolic blood pressure
GLP-1RA: glucagon-like peptide-1 receptor agonists
MD: mean difference
RCTs: randomized controlled trials
SBP: systolic blood pressure
T2DM: type 2 diabetes mellitus

## Statement of Ethics

Ethics approval for this paper is not required because this study is based exclusively on published literature. Patient consent was not needed as this study was based on publicly available data.

## Author Contributions

**Frederick B. Rivera MD:** Conceptualization; Data curation; Formal analysis; Investigation; Methodology; Validation; Writing - original draft; Writing - review & editing**; John Paul Aparece, MD, Grace Nooriza O. Lumbang, MD;Danielle Rose Magno Gaid, MD, Linnaeus Louisse A. Cruz, MD:** Writing – Data curation; review & editing; **Grace Nooriza O. Lumbang, MD;Danielle Rose Magno Gaid, MD**, **Frederick B. Rivera MD** and **Linnaeus Louisse A. Cruz, MD:** Data Curation; Writing - original draft; Writing - review & editing; **John Vincent Magalong, MD:** Writing - original draft; Writing - review & editing; **John Vincent Magalong, MD and Nathan Ross B. Bantayan, BSc:** Data curation; Data interpretation; Formal analysis; Writing - original draft; Writing - review & editing; **Frederick B. Rivera, MD:** Writing - original draft; Writing - review & editing; **Kyla M. Lara-Breitinger, MD; Martha Gulati, MD MS; George Bakris, MD**: Writing - review & editing.

## Data Availability Statement

All data generated or analyzed during this study are included in this article. Further inquiries can be directed to the corresponding author.

---

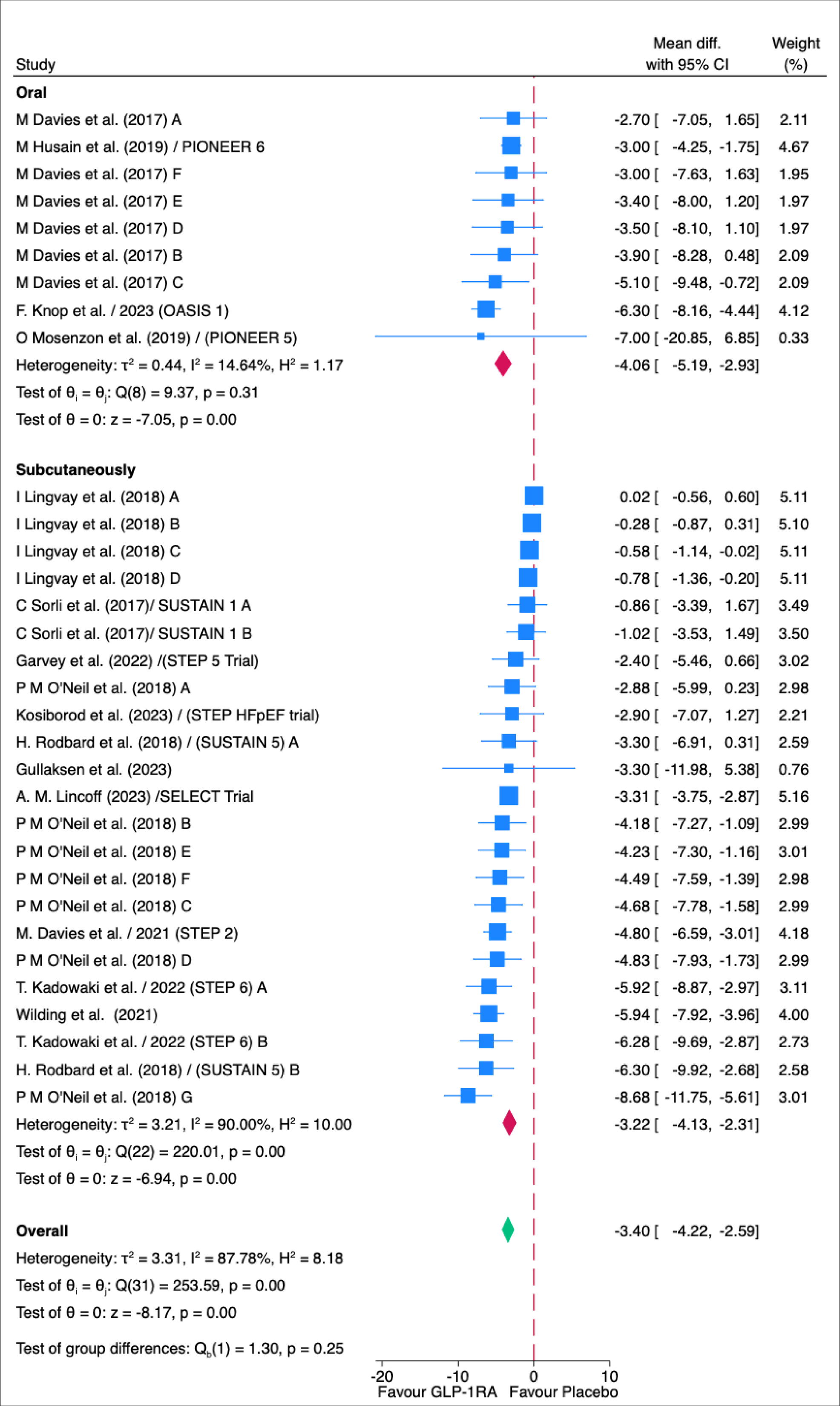

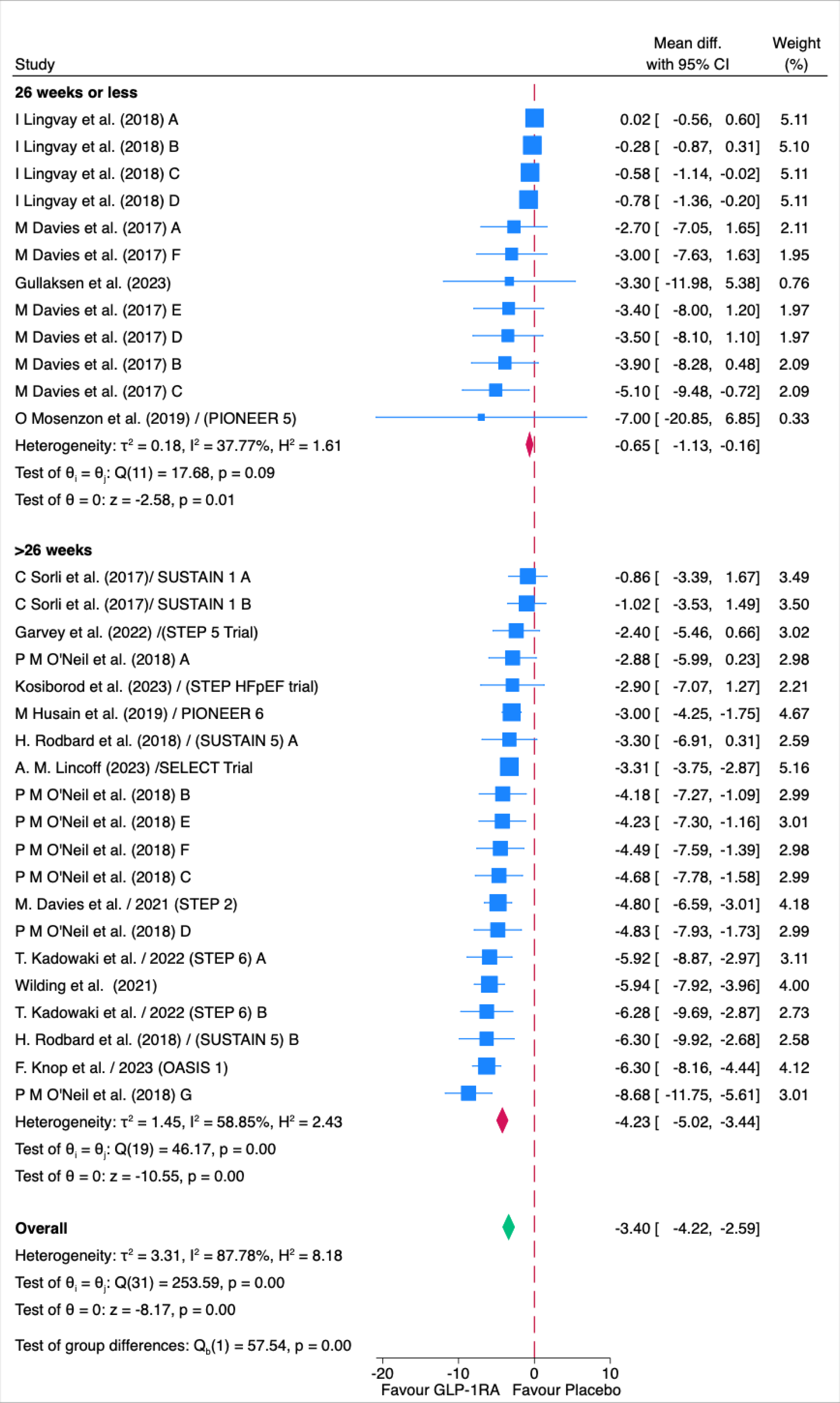

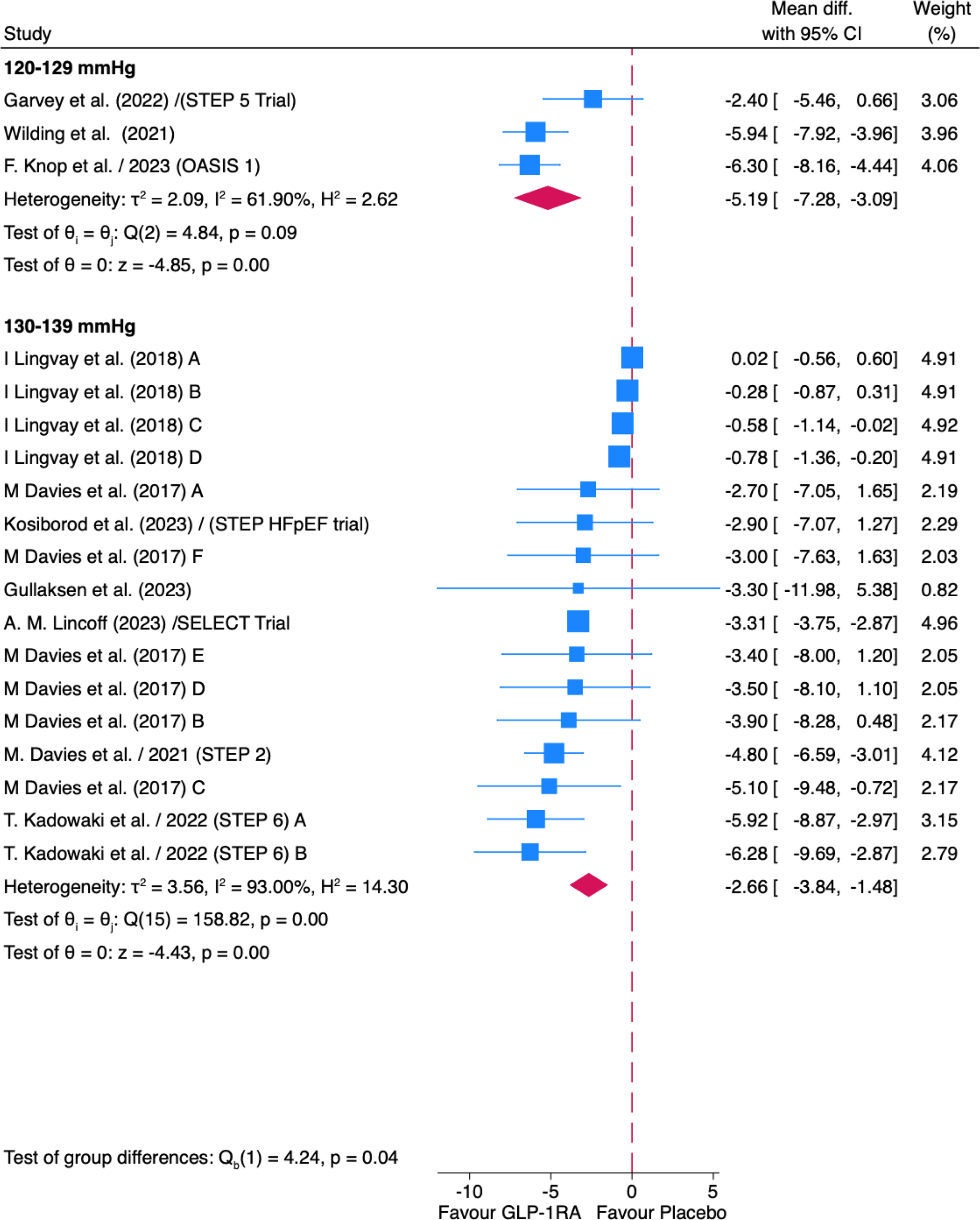

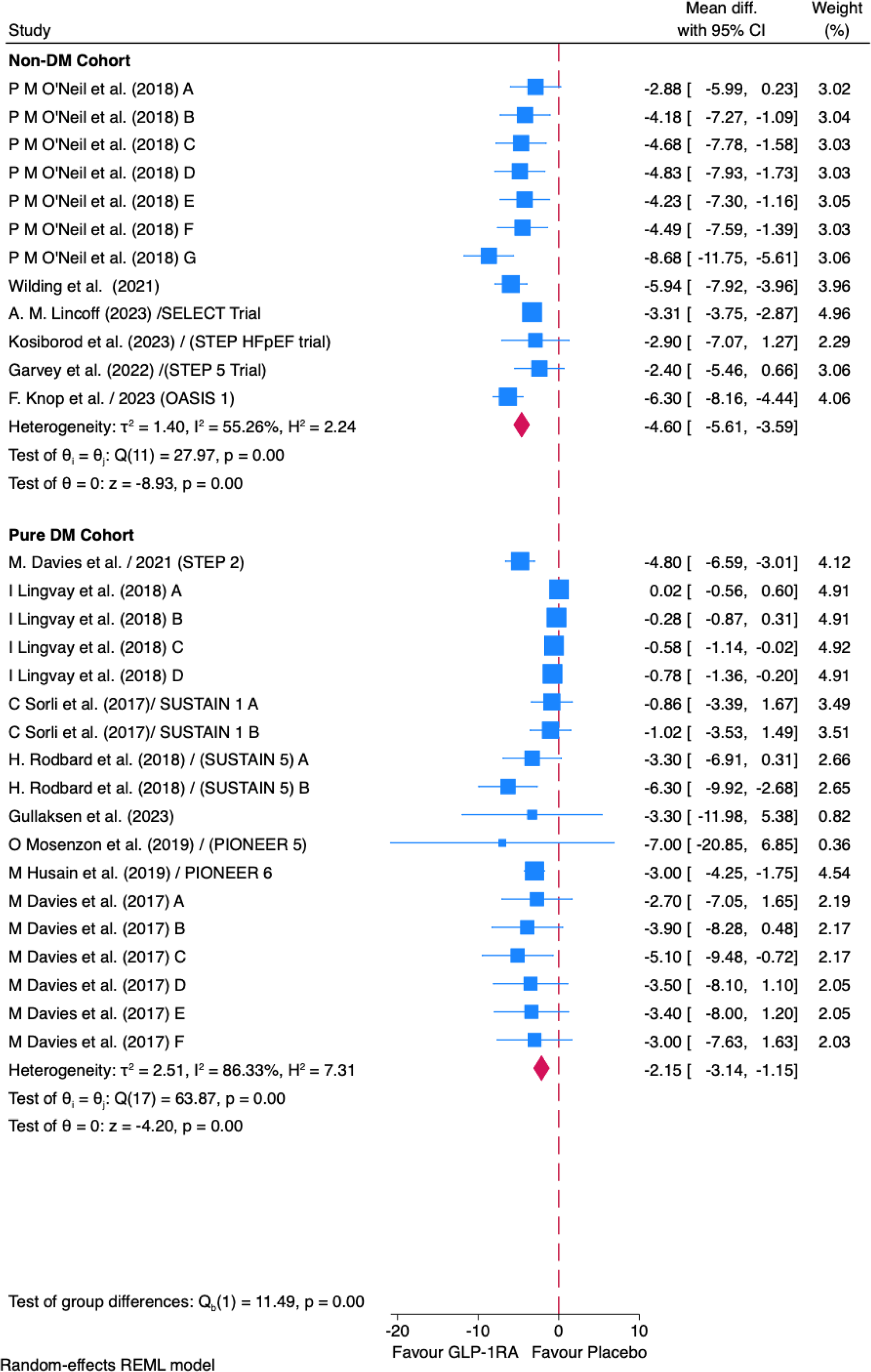

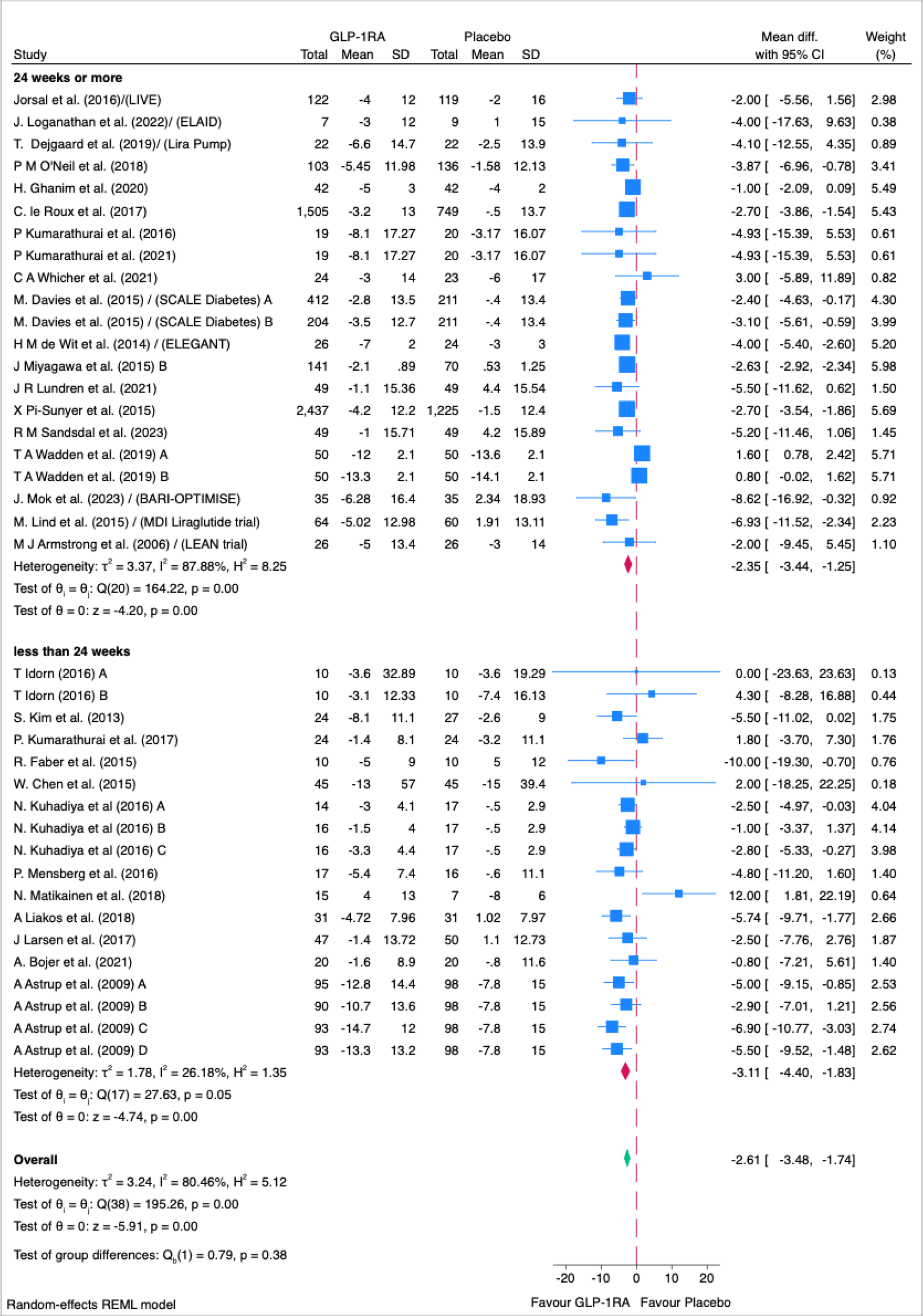

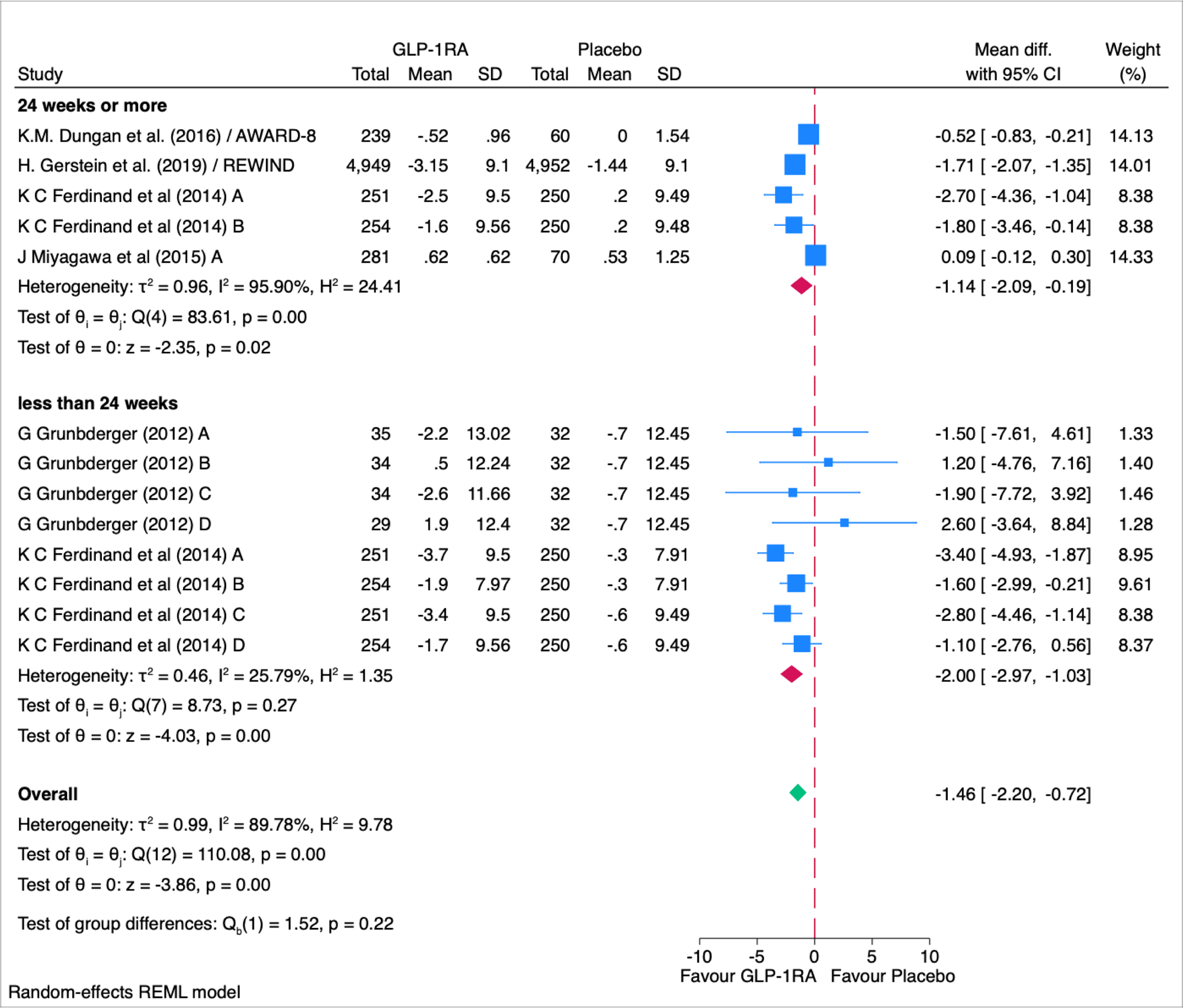

